# Unravelling the complexities of the first breaths of life

**DOI:** 10.1101/2020.07.29.20161166

**Authors:** David G Tingay, Olivia Farrell, Jessica Thomson, Elizabeth J Perkins, Prue M Pereira-Fantini, Andreas D Waldmann, Christoph Rüegger, Andy Adler, Peter G Davis, Inéz Frerichs

**Author notes:** Corresponding Author: A/Prof David G Tingay MB BS DCH FRACP Ph.D., Neonatal Research, Murdoch Children’s Research Institute, Royal Children’s Hospital Flemington Rd., Parkville 3052 Victoria, Australia. **Descriptor:** 14.3 Neonatal Lung Disease & BPD. This article has an online data supplement, which is accessible from this issue’s table of content online at www.atsjournals.org. **Author Contributions:** DGT developed the concept and designed the experiment. DGT, OF, JT, EJP, CR enrolled and studied all infants. AW, IF, AA, DGT developed the image reconstruction and analysis methods used in the study. DGT, OF, JT, PP-F were involved in data analysis. DGT, PGD, IF, AA interpreted the data. DGT wrote the first draft of the manuscript and all authors contributed to redrafting the manuscript. **Competing Interests:** AW was an employee of Swisstom AG (Landquart, Switzerland), who initially developed the EIT hardware and software systems (Swisstom was acquired by Sentec AG after the study was completed). Swisstom was not involved in study design, implementation, analysis, interpretation or reporting. IF and AW were investigators in an European Union Horizon 2020 Research and Innovation program grant to develop an infant EIT chest imaging system (CRADL project, Grant ID 668259). IF also reports funding from another European Union Horizon 2020 project (WELMO, Grant ID 825572) and reimbursement of soeaking fees, congress and travel costs by Dräger Medical (a company that produces a commercial EIT unit). No author received an honorarium, grant, or other form of payment to produce the manuscript. The study was not commissioned and no commercial agencies were involved in any aspect of this study. The authors have no other competing interests to declare. **Ethics Approval:** The Royal Women’s Hospital Human Research and Ethics Committee (#16-33), and registered with the Australian New Zealand Clinical Trials Registry (ACTRN12618000128291). **Data Sharing:** Individual participant data collected during the study, after de-identification, and study protocols and statistical analysis code are available beginning 3 months and ending 23 years following article publication to researchers who provide a methodological sound proposal, with approval by an independent review committee (“learned intermediatry”) identified for purpose. Data is available for analysis to achieve aims in the approved proposal. Proposals should be directed to; to gain access, data requestors will need to sign a data access or material transfer agreement approved by the Murdoch Children’s Research Institute.

## Abstract

**Rationale:** The transition to air-breathing at birth is a seminal respiratory event common to all humans, but the intrathoracic processes remain poorly understood.

**Objectives:** The objectives of this prospective, observational study were to describe the spatiotemporal gas flow, aeration and ventilation patterns within the lung in term neonates undergoing successful respiratory transition.

**Methods:** Electrical impedance tomography was used to image intrathoracic volume patterns for every breath until six minutes from birth in neonates born by elective cesearean section and not needing resuscitation. Breaths were classified by video data, and measures of lung aeration, tidal flow conditions and intrathoracic volume distribution calculated for each inflation.

**Measurements and Main results:** 1401 breaths from 17 neonates met all eligibility and data analysis criteria. Stable functional residual capacity was obtained by median (IQR) 43 (21, 77) breaths. Breathing patterns changed from predominantly crying (80·9% first minute) to tidal breathing (65·3% sixth minute). From birth tidal ventilation was not uniform with the lung, favouring the right and non-dependent regions; p<0·001 versus left and dependent (mixed effects model). Initial crying created a unique pattern with delayed mid-expiratory gas flow associated with intrathoracic volume redistribution (pendelluft flow) within the lung. This preserved functional residual, especially within the dorsal and right regions.

**Conclusions:** The commencement of air-breathing at birth generates unique flow and volume states associated with marked spatiotemporal ventilation inhomogeneity not seen elsewhere in respiratory physiology. At birth neonates innately brake expiratory flow to defend functional residual capacity gains and redistribute gas to less aerated regions.

**At a Glance Commentary:** *Scientific Knowledge on the Subject:* Birth requires the rapid transition from a fluid-filled to aerated lung that is poorly understood. Limited human and animal studies suggest high intrathoracic pressure and flow states are required to attain functional residual capacity and support tidal ventilation.

*What this Study Adds to the Field:* In the first breath-by-breath imaging of the lungs of term neonates undergoing successful respiratory transition at birth we identified highly inhomogeneous, spatiotemporal aeration and ventilation patterns during. Crying at birth preserved functional residual capacity by allowing intrathoracic volume redistribution (pendelluft flow) within the lung. Newborns defend aeration from intrathoracic lung-fluid shifts at birth by innately braking expiratory flow using the glottis and diaphragm.

## INTRODUCTION

The rapid adaptation to air-breathing at birth (aeration) is one of the most important, but least understood, physiological events in humans. Much of our understanding is inferred from preclinical studies (1-4) or invasive observational studies.(5-8) These studies suggest that creating a functional residual capacity (FRC) during the initial process of lung aeration requires first clearing the airways of fetal lung liquid using high intra-thoracic pressure gradients.(1,2,5) Subsequent tidal ventilation must prevent influx of fluid back into the alveoli during expiration.(1) Animal studies have demonstrated that these processes exhibit a high degree of spatiotemporal variability within the lung.(2,9) For most newborns, fluid clearance and the transition from placenta to lung as the organ of gas exchange is achieved through the spontaneous onset of breathing. When this process fails, especially in preterm infants, death or significant morbidity may result. Due to an inability to define the processes of aeration and ventilation at birth, effective evidence-based interventions to support breathing after birth are lacking.(10,11)

The development of effective delivery room interventions first requires an understanding of the physiological processes defining success or failure of aeration at birth. Adapting physiological concepts from preclinical studies have limited utility as instrumentation restricts the ability to emulate respiratory mechanics and the neurological state of the breathing human infant.(1-3,12) The delivery room further creates a challenging research environment, the time critical and dynamic nature of birth itself hampers physiological measurements.(13) Lung volume changes at birth have been intermittently imaged using chest radiography(14) and ultrasound(15), and pressure and flow patterns measured invasively at the mouth or pharynx.(5,16) These studies identified unique breath types associated with high intra-thoracic pressure gradients during successful respiratory transition in term infants, specifically crying and grunting.(5,17,18) Importantly, these studies failed to directly define the fundamental dynamic spatiotemporal processes of aeration and subsequent ventilation within the lung.

To address this gap in knowledge, we used electrical impedance tomography (EIT), an emerging radiation-free imaging modality.(19) EIT uses the differential electrical properties of aerated and fluid-containing tissue to measure the tidal and end-expiratory volume changes in lung regions within a transverse chest slice.(19) We adapted our EIT techniques for measuring the respiratory transition in preclinical studies.(2-4,12,20-23) This allowed non-invasive, and non-hazardous, direct imaging of the dynamic breath-to-breath regional process of aeration at birth in human infants without interfering with normal physiology or clinical care. The objective of this study was to describe the spatiotemporal respiratory patterns associated with the successful transition to air-breathing after birth in term infants. The specific aims were to 1) characterise the inspiratory and expiratory time and flow characteristics within the lung at birth, and 2) describe the resultant spatiotemporal ventilation and volume patterns by breath type and time.

## METHODS

A detailed methodology can be found in the online supplement. This prospective observational study was conducted at the Royal Women’s Hospital, Melbourne, Australia.

Infants were eligible for enrolment if they were delivered by elective caesarean section via spinal anaesthesia for non-fetal reasons at ≥36^+0^ weeks gestation, and written prospective parental consent obtained. Infants were not included if placement of an EIT belt would interfere with clinical care,(24-26) or the fetus had a known congenital condition that would alter EIT interpretability. Infants who received resuscitative interventions were excluded from analysis.

### Measurements

Heart rate and peripheral oxygen saturation (SpO_2_) were measured with a Radical 7 pulse oximeter (Massimo Corporation, Irvine CA). Regional lung volume changes were imaged at 48 frames/s with the Pioneer EIT system using an ultrasound gel-coated NeoSensor Belt (Sentec AG, Landquart, Switzerland).(2,12,24-26) Audio and video were recorded at 30 frames/s (Logitech webcam, Lausanne, Switzerland).

### Delivery room protocol

As the infant was being placed supine on the resuscitaire the NeoSensor Belt was secured (velcro tab) around the chest at nipple level (**Supplementary Video 1**). The pulse oximetry sensor was applied to the right hand. There was no other interference with routine clinical care. Infants were managed in a supine position in accordance with local guidelines, including timing of umbilical cord clamping. Data were only recorded during care on the resuscitaire.

### Data acquisition and analysis

EIT, video, audio and pulse oximetry data were continuously recorded digitally during resuscitaire management, and timing of critical events from birth documented. SpO_2_ and heart rate data were reviewed for loss of signal or movement artefact. EIT data were recorded in a custom-built infant imaging package,(27) and images reconstructed post hoc(19,28) using the vendor-provided human model chest atlas, with non-lung regions excluded.(2,23,24,26) Each potential tidal volume (V_T_) change due to breathing was identified from the global lung signal. Analysis of the EIT change associated with a breath was only performed if there was 1) video confirmation of a breath; and 2) no movement interference on the video. All included breaths were classified by the presence of an audible cry, grunt or no breathing noise (tidal breath). If audio classification was not possible the breath was excluded (Supplementary Figure 1).

**Figure 1.**
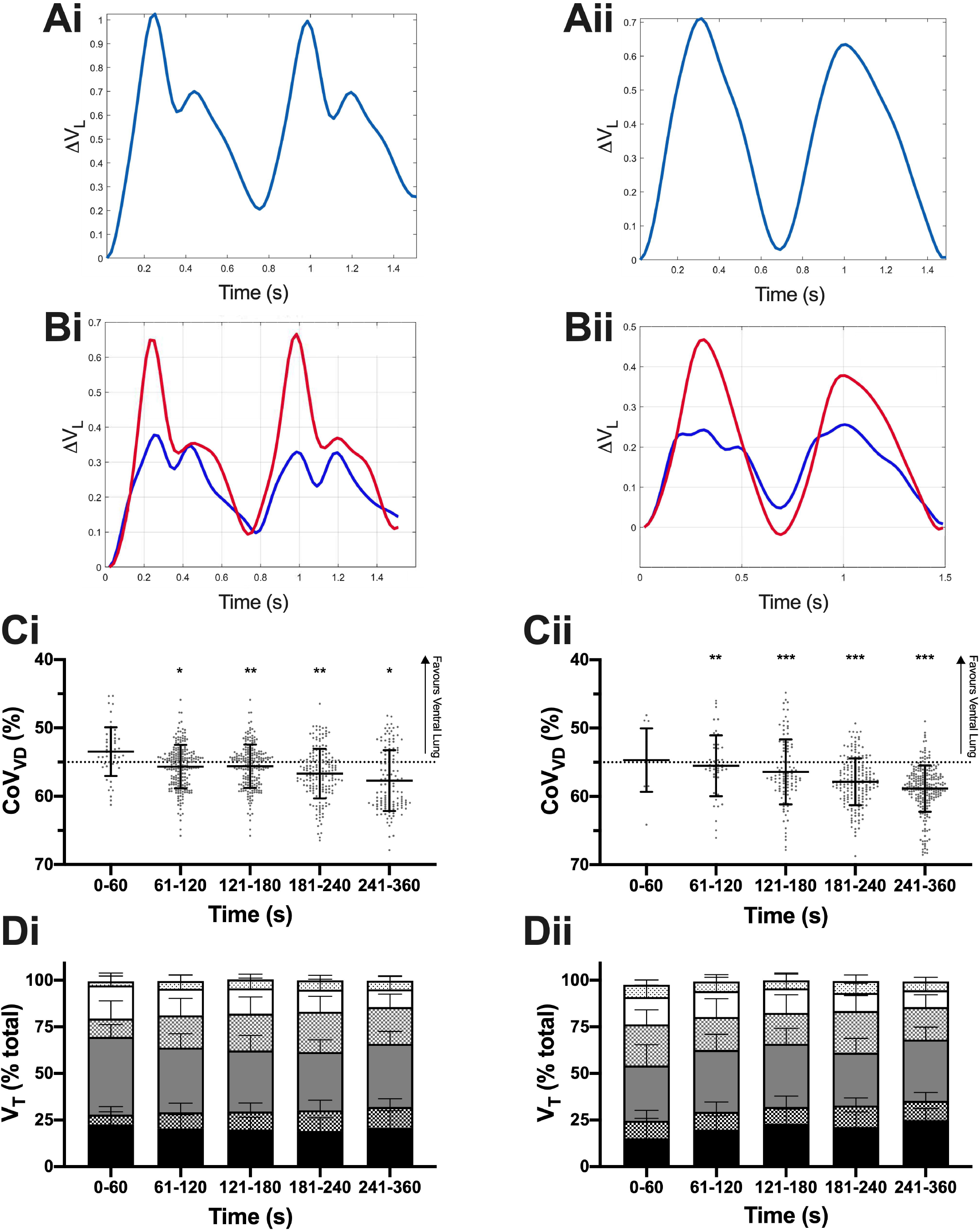
**A**. Relative Volume change (ΔV_L_) over time within the whole lung during the two representative breaths for crying (**i**; 25 s) and tidal breaths (**ii**; 5 min). **B**. ΔV_L_ during the same breaths within the ventral (blue) and dorsal (red) hemithoraces. **C**. Ventrodorsal centre of ventilation (CoV_VD_) by minute after birth for crying and tidal breaths. CoV_VD_ of 55% represents uniform ventilation, with values <55% indicating relatively greater ventilation in the ventral lung and >55% the dorsal lung. Grey dots represent individual breath data, and black line and bars mean±SD. **C**. Relative distribution of ventilation (% total V_T_) along the gravity dependent plane for crying and tidal breaths in the most gravity dependent third of the lung (black bars), central third (grey bars) and non-gravity dependent third (white bars), with solid bars being right lung region and dotted bars left lung. All data mean+SD. *p<0.05,**p<0.01***p<0.0001 against first 60s (mixed-effects model).

For included breaths, the pre and post-breath FRC, inspiratory time (Ti), expiratory time (Te), time constant of the respiratory cycle (τ) and relative peak inspiratory (PIF) and expiratory flow (PEF) were calculated for the global signal and right, left, ventral and dorsal lung regions.(19) The shape of the impedance change during each breath was classified by an investigator (DGT) unaware of the breath type or time from birth. The centre of ventilation along the ventrodorsal (CoV_VD_) and right-left (CoV_RL_) planes were calculated to determine spatiotemporal distribution of V_T_ within the chest slice.(19,29) The percentage of the global V_T_ signal was calculated for the most dependent, central and non-dependent thirds of the right and left lung regions, and the percentage and location of lung regions without any V_T_ signal.(19,24)

### Sample size and statistical analysis

Based on a previous study of respiratory parameters at birth (16), a convenience sample of thirty infants was estimated to provide data for breath-by-breath classification. and analysis of 100-150 breaths/infant per 5-6 minute period in 15-20 infants. Continuous data were analysed with a mixed-effects linear regression model, with robust standard error and cluster analysis to adjust for multiple breaths from each infant. A p value <0.05 was considered statistically significant.

## RESULTS

A visual abstract of the main study findings is available in **Supplementary Video 2**.

### Study population

Thirty-three families were approached on the day of the delivery, with three declining to participate. Two studied infants received resuscitative support after birth and were excluded. Of the remaining 28 infants, EIT data were obtained in 27 infants (EIT belt incorrectly placed). Complete audio or video data were not acquired in ten infants (technical failure in delivery room, camera obstructed, excessive background noise or inability to delineate any audio/video breaths). The characteristics of the final 17 infants with matched EIT, video and audio data are described in Supplemental Table 1. All were singleton pregnancies and no mother received antenatal corticosteroids.

**Table 1.**
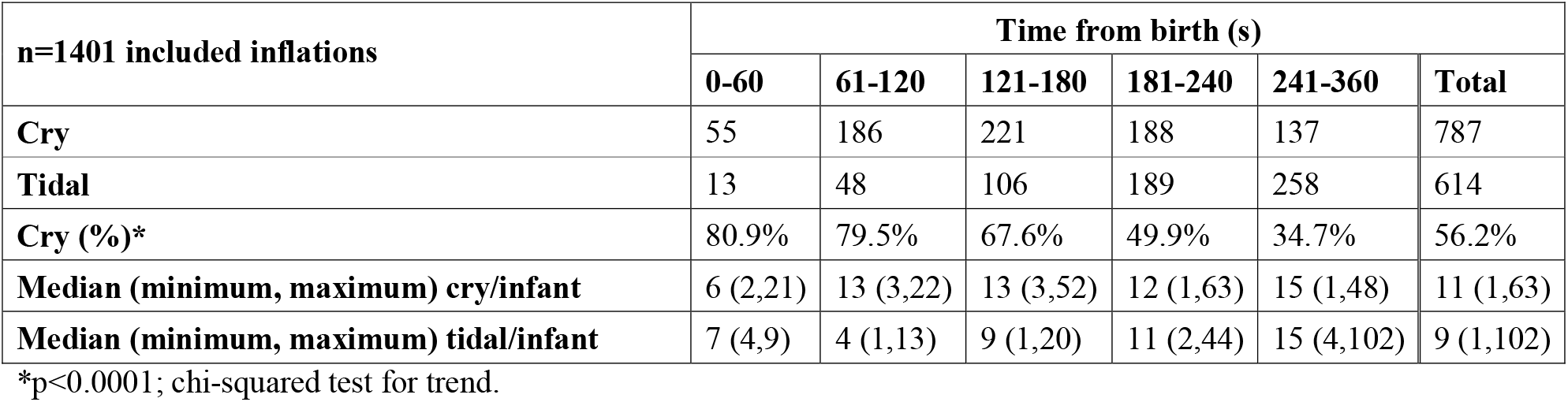
Type of breath by time.

| **n=1401 included inflations** | **Time from birth (s)** |  |  |  |  |  |
| --- | --- | --- | --- | --- | --- | --- |
|  | **0-60** | **61-120** | **121-180** | **181-240** | **241-360** | **Total** |
| **Cry** | 55 | 186 | 221 | 188 | 137 | 787 |
| **Tidal** | 13 | 48 | 106 | 189 | 258 | 614 |
| **Cry (%)\*** | 80.9% | 79.5% | 67.6% | 49.9% | 34.7% | 56.2% |
| **Median (minimum, maximum) cry/infant** | 6 (2,21) | 13 (3,22) | 13 (3,52) | 12 (1,63) | 15 (1,48) | 11 (1,63) |
| **Median (minimum, maximum) tidal/infant** | 7 (4,9) | 4 (1,13) | 9 (1,20) | 11 (2,44) | 15 (4,102) | 9 (1,102) |

\*p<0.0001; chi-squared test for trend.

### Pulse oximetry

A pulse oximetry signal could be acquired in 15 infants, with a median (range) of 17 (3, 198)s between applying the probe and signal acquisition. The first SpO_2_ signal was acquired at 52 (12, 97)s, but then lost for >10s at least once in ten infants. SpO_2_ increased with time from 53 (48, 72)% at 60s to 78 (60, 96)% by 360s (p=0.029; mixed-effects model, Supplementary Figure 2). Heart rate was stable throughout the study period (p=0.25).

**Figure 2.**
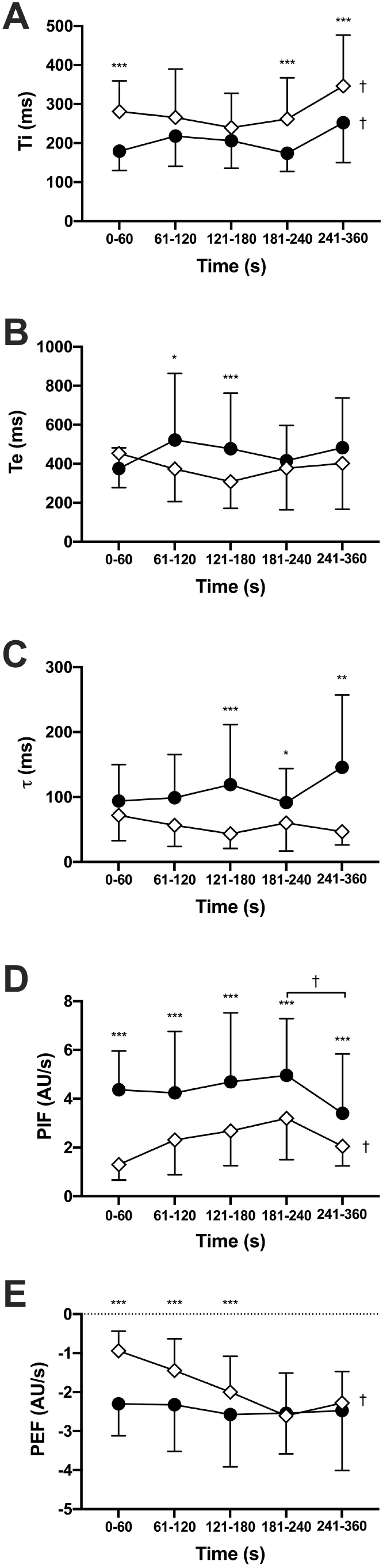
Ti (**A**), Te (**B**), respiratory system time constant (τ; **C**), PIF (**D**) and PEF (**E**). Black circles indicate crying breaths and white diamonds tidal breaths. All data mean±SD. *p<0.05, **p<0.01, ***p<0.0001 cry vs tidal inflation; †p<0.01 within breath type (all mixed-effects model).

### Time to image acquisition

The median (range) time from birth to first EIT image was 36 (20, 62)s, with the longest periods being in the two infants born with delayed cord clamping (60 and 62s). The time between cutting of umbilical cord and first EIT image was 31 (20, 48)s. In all infants the time between applying the EIT belt and first images was <12 s, with no subsequent signal loss.

### Breathing patterns

A total of 1401 inflations met the inclusion criteria (Table 1). Only 14 breaths (1%) had audible grunting (all during periods of crying), and were included within the 787 crying breaths. Overall, crying was more prominent early in the respiratory transition, representing 80.9% of all included inflations within the first minute, then decreasing to 34.7% by the sixth minute (p<0.0001; chi-squared test for trend).

Breaths could be classified as following two distinct EIT volume patterns; 1) linear inspiratory and expiratory volume change consistent with tidal ventilation of already-aerated lungs, or 2) an expiratory phase with a distinct bifid expiratory wave and a transient increase or preservation in lung volume (Figure 1 and **Supplementary Video 2**). During these bifid waves there was a subtle redistribution of ventilation seen on fEIT images consistent with pendelluft flow. 70.8% of all crying breaths had a bifid wave, but only 2.5% of tidal breaths.

Ti increased over the first minutes of life for both breath types; p<0.0001 (Figure 2A). Crying generated shorter Ti than tidal breaths in the first 60s, mean (95% CI) difference 101 (56, 147) ms. In contrast, Te and τ were longer during crying than tidal breaths (both p<0.0001; Figure 2B and C), especially between 61-180s (Te) and after 120s (τ). Overall, Te and τ did not change significantly with time for crying or tidal breaths.

PIF was greater at all time epochs during crying compared to tidal breaths (all p<0.0001). Overall, PIF increased with time for tidal breaths (p<0.0001; Figure 2D), but not during crying. PEF was greater during crying than tdal breaths for the first 180s (all p<0.0001; Figure 2E), with the greatest difference in the first 60s (1.4 (0.9, 1.8) AU/s). PEF decreased with with time during crying (p<0.0001), whilst tidal breaths were unchanged.

The detailed spatiotemporal behaviour of Ti, Te, τ, PIF and PEF in the right, left, ventral and dorsal regions are provided in the supplemental results (Supplemental Figures 3-7). Overall, Ti, Te and τ were similar within all regions for both breath types. Crying resulted in faster PIF and PEF in the dorsal and right regions compared to ventral and left respectively. Tidal breaths resulted in less right-left and ventral-dorsal heterogeneity in PIF and PEF than crying.

**Figure 3.**
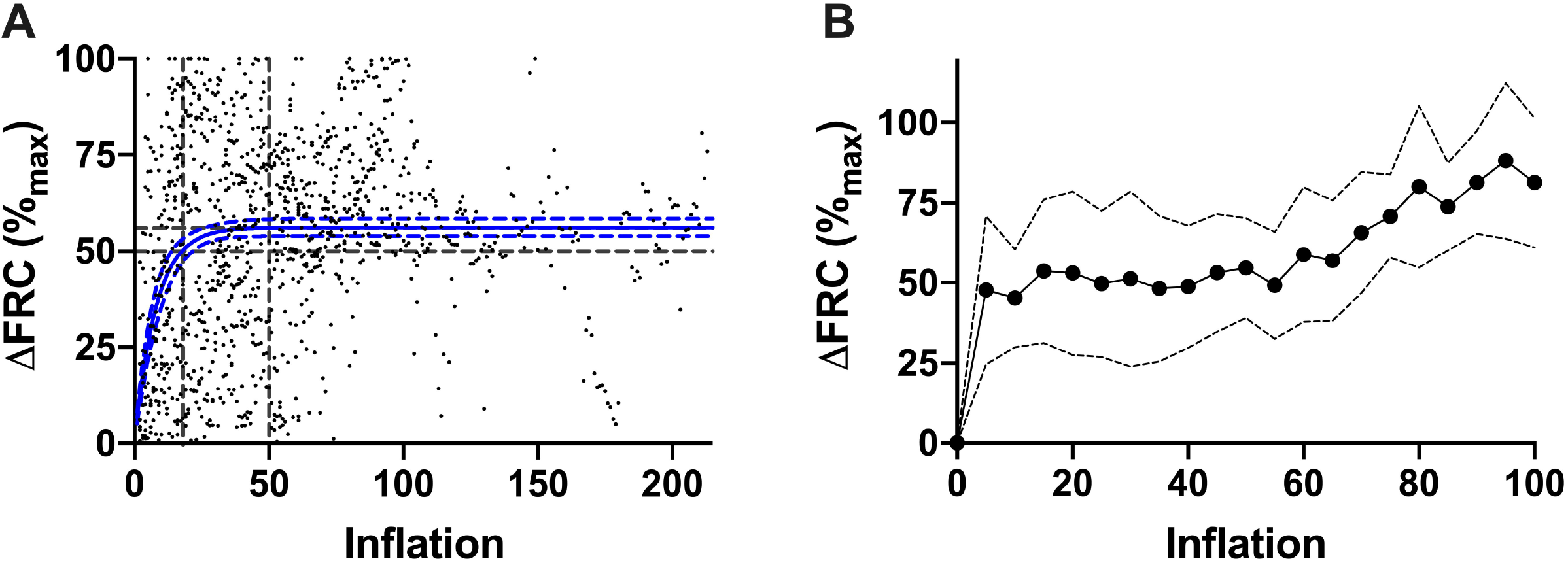
**A**. Change (Δ) in FRC from first measured inflation for all breaths (**A**) and for the first 100 inflations (maximum)(**B**). ΔFRC normalised to the FRC before the first breath (0%) and maximum FRC (100%) for each infant. **A**. Blue line represents line of best fit (dashed lines 95% CI) using a one-phase exponential association; y=y_plateau_.[1-*e*^.τ-1^]; plateau (95% CI) 56.2 (54.0, 58.6)%, τ 8.0 (6.0, 10.5) inflations (R^2^ 0.14, RSME 24.5%, replicates test discrepancy (F) 0.82 [p=0.96]). Grey dashed lines demonstrate ΔFRC at 50% of FRC_max_ and 50 inflations. **B**. Black circles represent the mean ΔFRC every 5 inflations for each infant, and dashed lines 95% CI.

### Functional residual capacity

Overall, FRC increased, and was quickly established, after birth, with the maximum recorded FRC value for each infant occurring at a median (IQR) 43 (21, 77) of included breaths after birth; 67.8 (51.9, 94.5)% of the analysed sequential breaths (Figure 3 and Supplemental Figure 8). During an infant’s first 100 breaths (or total if <100), 48% of FRC change occurred by the fifth breath.

### Regional ventilation patterns

Ventilation redistributed towards the dorsal regions with time for both crying and tidal breaths (Figure 1); CoV_VD_ p=0.045 and p<0.0001 respectively. Overall, CoV_VD_ favoured the ventral regions during crying compared to tidal breaths by a mean (95% CI) 1.6 (0.3, 2.9)%, although the differences were not significant within each minute. The redistribution of V_T_ towards the dorsal regions was predominantly due to increased V_T_ within central lregions during crying, and the dorsal region during tidal breaths (p=0.0004), both at the expense of ventral V_T_.

Both breath types resulted in greater ventilation in the right lung (Figure 4). During crying a mean (SD) 82.2 (15.3)% of total V_T_ occurring in the right lung during the first 60s (CoV_RL_ 3.0 (9.1)%; ideal 46%), increasing within the left lung with time (p=0.011). By 240s 64.3 (11.4)% of V_T_occurred within the right lung (CoV_RL_ 43.1 (6.5)%). The right lung accounted for predominance of 59.5 (14.3)% of V_T_ (CoV_RL_ 46.4 (9.3)%) during tidal breaths within the first 60s, and this predominance did not change over time (p=0.10). Crying resulted in greater right-left lung inhomogeneity, with CoV_RL_ being a mean (95% CI) 2.0 (0.5, 3.5)% less overall than tidal breaths, and the difference greatest in the first minute; 13.4 (6.7, 20.1)%.

**Figure 4.**
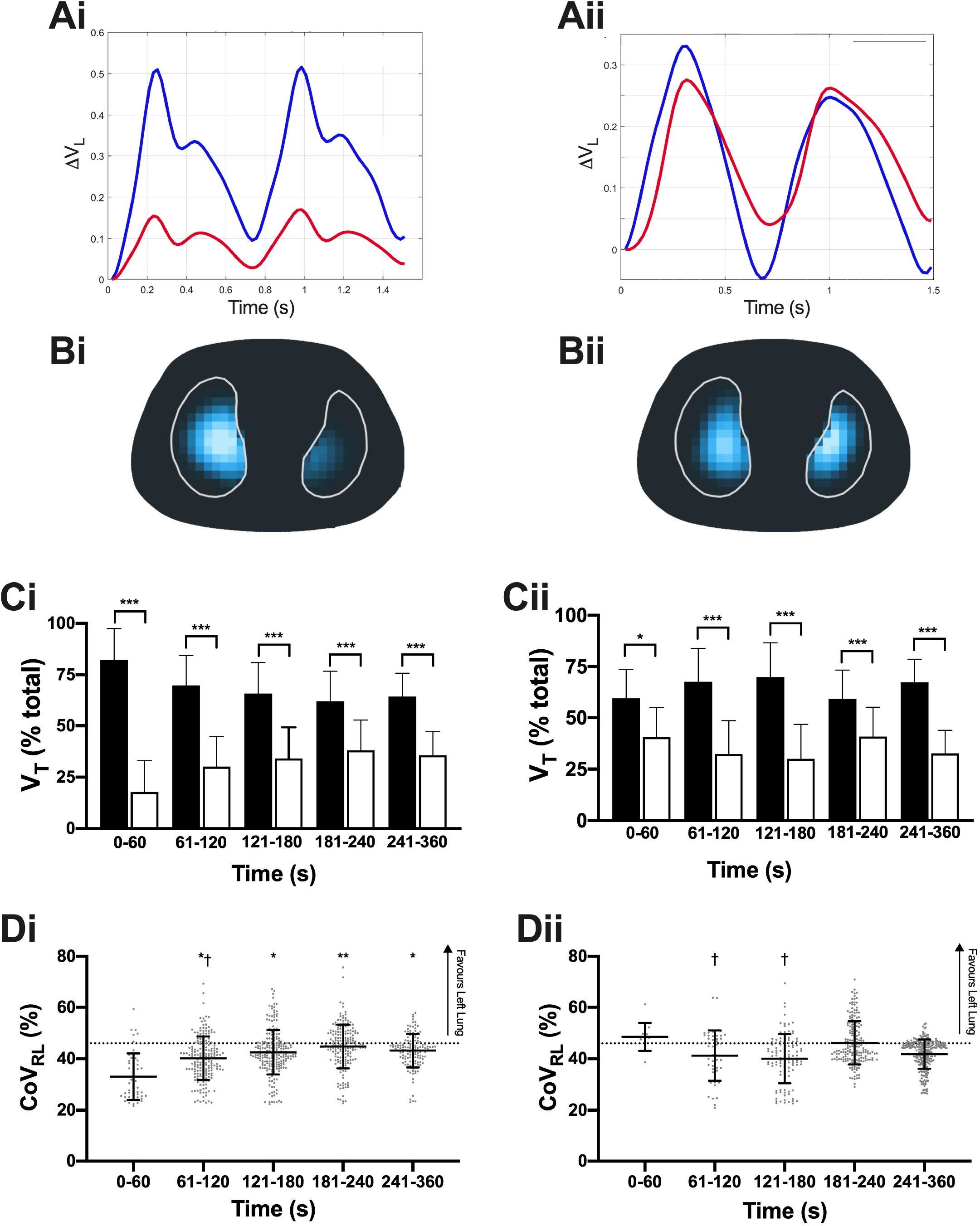
**A**. Relative Volume change (ΔV_L_) over time within the right (blue) and left (red) lung during the two representative breaths for crying (**i**; 25 s) and tidal breaths (**ii**; 5 min). **B**. Functional EIT images of volume change within the lungs for the same breaths using the colour scale defined in Supplementary Video 2. **C**. Relative distribution of ventilation (% total V_T_) in the right (black bars) and left (white bars) lung for crying and tidal breaths. **D**. Centre of ventilation (CoV_RL_) along the right-left plane for all inflations by minute after birth for crying and tidal breaths. CoV_RL_ of 46% represents uniform ventilation, with values <46% indicating relatively greater ventilation in the right lung, and >46% greater ventilation in the left lung. All data mean±SD, and dots individual breath data. Panel A and C; *p<0.05, ***p<0.0001 (mixed-effects model). Panel B and D; *p<0.05, **p<0.01 against first 60s. † p<0.05 against 181-240s.

Approximately 10% of predefined lung regions were unventilated for both tidal and crying breaths, with not difference in the ventrodorsal pattern of unventilated regions (Supplementary Figure 9). After 240s there was less unventilated lung regions, especially during tidal breathing, suggesting increasing aeration resulted in greater engagement of the distal lung in ventilation.

## DISCUSSION

The transition to air-breathing at birth is a seminal physiological event essential to life in all humans. In our observational study we provide the first detailed description of the volumetric processes within the lung at birth. We found that the transition to air-breathing is characterised by complex spatiotemporal patterns of aeration and ventilation initially mediated by high PIF rates and prolonged expiration. Overall this results in rapid lung aeration that moves from the central to distal lung, with the right lung engaging in ventilation earlier than the left. Crying, the dominant breathing pattern at birth, creates greater PIF and complex expiratory volume patterns, including pendulluft flows, more suited to both rapid aeration and maintenance of FRC than tidal breathing, at a time the lung is still likely to be partially fluid-filled. That these findings occurred in healthy term infants without instrumentation or active intervention is important, providing the first human evidence that successful aeration at birth is dependent on actively engaging in expiratory mechanisms to protect FRC (**Figure 5**).

**Figure 5.**
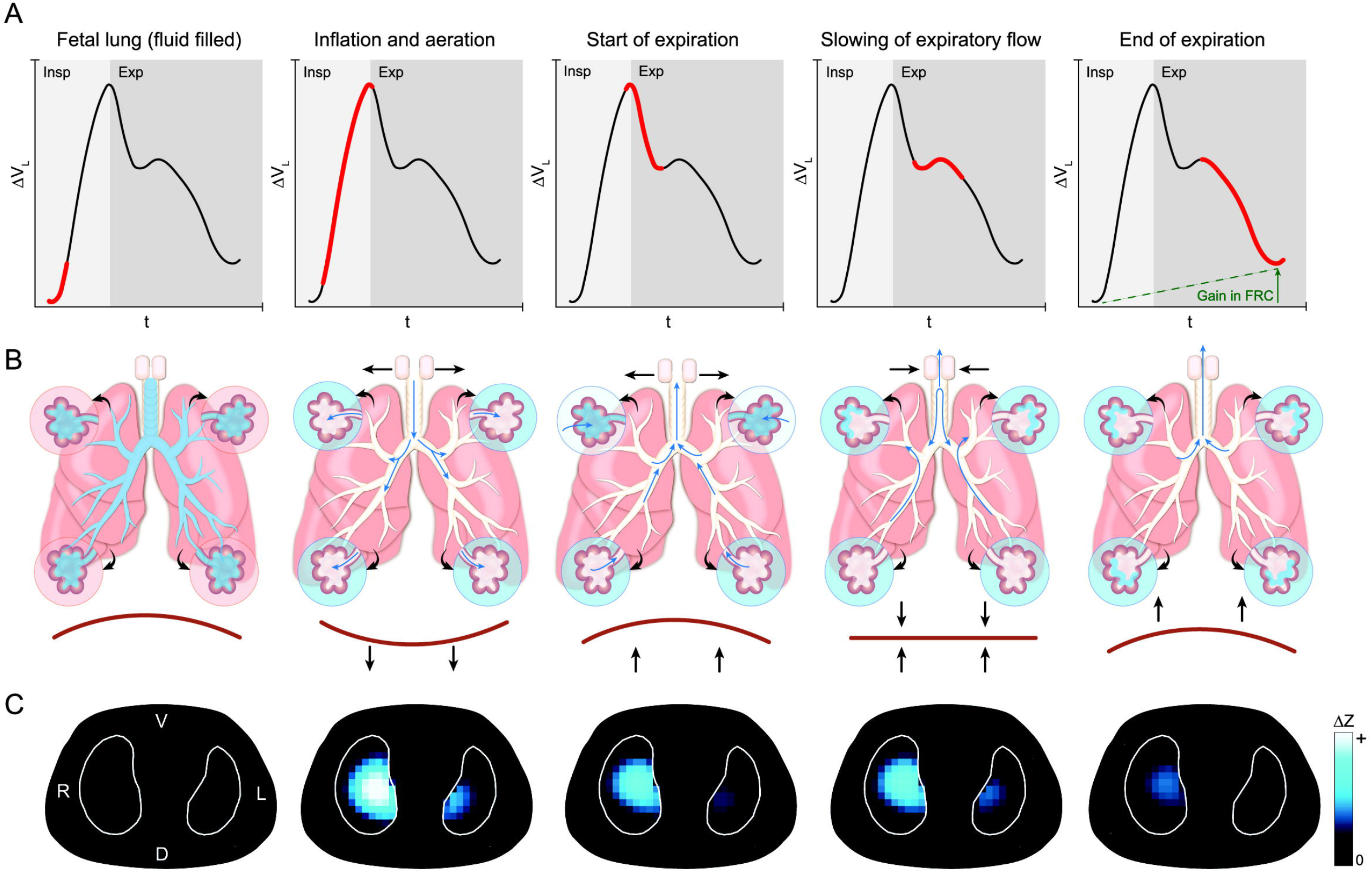
Summary of main findings and hypothesed explaination of the criteria which define the respiratory events within the lungs at birth. Representative global lung volume change during a single cry at 14 s after birth (Infant 6; **Panel A**) demonstrating the dynamic volume change during inspiration and expiration, and resultant increase in end-expiratory FRC. The breath has been divided into five phases related to the mechanistic events identified during the respiratory transition from a fluid-filled to aerated lung (**Panel B**), and the respective fEIT images for each shown in **Panel C**. At the start of the inflation the airways and alveoli are fluid filled **(Column 1**). A cry initiates a rapid and large contraction of the diaphragm with a resultant rapid inspiratory flow (slope of the time-volume curve) and high inflating (driving) pressure within the lung generating aeration by moving fluid from the proximal airways to the alveoli (enlarged) and then the lung interstitium (**Column 2**). Expiration begins with rapid contraction of the diaphragm **(Column 3**). The fall in intra-thoracic pressure during expiration lowers intra-alveolar pressure, and in some lung units this fall allows fetal fluid to influx back into the alveoli spaces. To counteract this effect the neonate slows (brakes) diaphragmatic contraction and partially closes the glottis, thus transiently re-pressurising the lung and allowing pendulluft gas flow between aerated to poorly aerated lung units (**Column 4**). When expiration continues it does so against a partially closed glottis which mediates slower expiratory gas flow and allows some gas to remain in the lungs, thus generating a greater end-expiration FRC **(Column 5**) and more favourable lung conditions at the start of the next inflation.

Clearing the respiratory system of fetal lung liquid, and establishing aeration is essential to physiological success at birth. We showed that the majority of lung aeration is rapidly achieved at birth, similar to chest radiography studies during the first seconds after birth in term infants.(7,8) Unlike these studies we were able to continuously follow the process of aeration beyond the first inflations. Although there was considerable inter-subject variability, aeration conformed with an exponential pattern reported in preclinical studies,(3,4,23,30,31) and during lung recruitment in the already-aerated lung.(32,33) Aeration was also associated with a temporal increase in distal lung ventilation. Our study is the first in humans to confirm the sequential central-distal movement of the air-fluid interface during aeration from the major airways to the distal alveoli reported in animal studies is ongoing beyond the first few breaths.(1,9) EIT can not directly measure airspace fluid, but the markedly different electrical properties of air and fluid make EIT ideally suited to mapping the air-fluid interface and tracking lung aeration clinically.

The patterns of ventilation indicate that spatiotemporal aeration after birth is more complex than only a central-distal process. The preferential ventilation of the right lung was unexpected but biologically plausible. At birth the lung is fluid filled, and airways (and tissue) have a high resistance.(34) The left main bronchus exits the carina acutely, and is encumbered by the heart. This may create preferential flow states towards the right lung, especially during the higher inspiratory flows of crying. Resistance falls in those areas of the lung that aerate first, further potentiating ventilation compared to unaerated regions. Our data also suggests that ventilation initially follows a gravity-dependent pattern, similar to that seen in parenchymal lung diseases.(35,36) Once aerated the lung rapidly develops the anatomical ventrodorsal pattern of ventilation reported in healthy older infants, favouring the dorsal lung with its increased lung mass and greater diaphragmatic tidal movement.(19,37) These changing spatiotemporal patterns across multiple planes make applying respiratory support without risking lung injury particularly challenging.

As expected in healthy infants, during the first two minutes 80% of breaths were cries. In a similar population of 13 infants, 77% of the analysed 749 breaths within the first 90 seconds after birth were classified as cries or grunts, but breath classification was performed post hoc from face mask measurements without auditory or visual confirmation, limiting interpretability.(16) Our study is the first to classify volume changes with flow and breathing behaviour. Crying created different flow characteristics than tidal breathing, quickly inflating the lung with faster Ti and PIF. This is advantageous within the highly resistive fluid-filled lung at birth,(34) but once aerated provides little mechanical or gas exchange benefit. We postulate that crying has a de novo physiological purpose, and not simply due to the noxious stress response of birth. Once aerated the high PIF conditions of crying increase unventilated lung tissue, with infants switching to the more advantageous tidal breaths.

Importantly, crying is also an expiratory phenomenon, being associated with slow expiratory flows, longer Te and τ. Volume loss during expiration followed a unique bifid pattern, occurring in 71% of all cries and rarely in tidal inflations. This pattern of volume change represents transient periods of minimal airway flow despite the chest wall being in a state of expiratory recoil. In this state reducing expiratory flow could only be achieved via active means, such as glottic closure or diaphragmatic hold; both seen in radiological imaging at birth.(8) The lung is in a state of flux in early ex-utero life; the alveoli maybe air-filled but fetal lung fluid remains in the interstitium, and fluid can influx back into alveoli if the intrathoracic pressure gradient falls, compromising FRC.(4,9,34) It has been proposed that ‘expiratory braking’ is essential during this period,(1,5,7,8,17,18,38) and flow patterns measured at the airway opening support this, but have not been correlated to temporal FRC change.(16) Our study provides the first evidence that expiratory braking does more than just prevent egress of gas from the lungs. It also facilitates the volumetric conditions needed to preserve FRC, and importantly redistributes gas within the lungs (pendelluft flow). We propose that this provides a simple visual indicator of an infant’s independent ability to independently support respiratory transition. Further studies are warranted to determine if the same breath types, and patterns, are present in at risk and preterm infants.

Reports of the cardiorespiratory processes at birth are sparse, mainly due to challenges in measurement. Following chest radiology studies in the 1960s,(7,8) instrumentation within the mouth, initially with bulky equipment,(5,17,18,38) and more recently, face masks(16,39,40) have measured airway opening flow, V_T_, expired CO_2_ and/or pressure changes, and infer intrathoracic conditions. Face masks are frequently applied with a leak,(41,42) application interferes with normal breathing efforts,(43) and cannot identify important spatiotemporal events, limiting usefulness during spontaneous breathing. Ideally measurements should be obtained from the thorax without impacting respiratory effort. Recently respiratory inductive plethysmography(39) and lung ultrasound(15) have been used in the delivery room. Inductive plethysmography requires two belts and determines lung volume from measuring the cross-sectional areas of the chest and abdomen, which may not change between fluid- and air-filled states.(32) Lung ultrasound is ideal for imaging the air-fluid interface, is simple to use, but lacks regional resolution, and continuous imaging has not been possible.(15) In this context, EIT is attractive. EIT is an established and validated method of measuring relative change in multiple spatiotemporal respiratory parameters.(19) EIT is radiation-free and available with a simple non-invasive belt (25) that could be applied as quickly, and more reliably, than pulse oximetry. EIT also confirmed the physiological patterns seen in humans and preclinical studies using these other measurement tools.(2,9,23) We contend that EIT is currently the best method of monitoring the respiratory system at birth.

### Limitations

Our study was limited to birth via elective caesarean section, and measuring were not made from delivery of the chest. The birth experience is uniquely personal, and we intentionally limited our study to a period of clinical mother-baby separation. Consequently, we missed the first few inflations in most infants. We content that these inflations are unlikely to be markedly different from those we captured. The unique volumetric, flow and FRC characteristics we identified indicate fluid clearance was still ongoing during the first 120s. It is unlikely that the enhanced lung liquid clearance provided by delivery through the vaginal canal would alter the respiratory findings in our healthy term population with active vigorous breathing. Respiratory effort was not suppressed, but data on all modes of delivery are needed in less vigorous infants. We have demonstrated that EIT can be practically applied earlier and during vaginal delivery. Our study of 1401 inflations from 17 infants is one of the largest, but, like previous studies,(16,17) exclusions were necessary and may have included potentially important breaths. In part this was intentional; our methodology was designed to minimise artefact and ensure correct breath classification lacking in previous studies. It is possible that respiratory drive occurred with an occluded airway. This would not result in a volume change on EIT but is an important physiological finding that should be seen on video. Like all other imaging tools used to describe the respiratory transition, EIT is limited to a single slice of the lung. However, single slice EIT has been shown to represent whole lung patterns in infants.(19) EIT cannot measure intrathoracic pressure. To do so would require invasive instrumentation, but is unnecessary as flow patterns reflect intrathoracic pressure states.

## Conclusions

This study provides the first detailed description of the respiratory behaviour of the healthy human lung during the transition to air-breathing after birth. Birth requires rapid aeration of the lung, and is achieved predominately via crying. Crying creates unique flow and volume states not seen elsewhere in respiratory physiology, and is characterised by high peak inspiratory flow and expiratory braking to preserve attained FRC and allow volume redistribution. The right lung ventilates before the left lung after birth, and the lung quickly develops an anatomical pattern of ventrodorsal ventilation once aerated. Understanding how the human lung successfully commences breathing at birth is the first step in developing tools to identifying when intervention is required.

## Supporting information

OSM Video 1

OSM Video 2

## Data Availability

Individual participant data collected during the study, after de-identification, and study protocols and statistical analysis code are available beginning 3 months and ending 23 years following article publication to researchers who provide a methodological sound proposal, with approval by an independent review committee (learned intermediatry) identified for purpose. Data is available for analysis to achieve aims in the approved proposal. Proposals should be directed to; to gain access, data requestors will need to sign a data access or material transfer agreement approved by the Murdoch Childrens Research Institute.

## ABBREVIATIONS

AU: Arbitrary Units
CoV: Centre of Ventilation
EIT: Electrical Impedance Tomography
FRC: Functional Residual Capacity
PEF: Peak Expiratory Flow
PIF: Peak Inspiratory Flow
SpO_2_: Peripheral oxygen saturation
τ: Time constant (tau)
Ti: Inspiratory time
Te: Expiratory time
V_T_: Tidal Volume

## Acknowledgements

The authors wish to thank the families, infants and Royal Women’s Hospital staff involved in the study. The authors acknowledge Dr Kate Patterson of Medipics and Prose for assistance with figures and videos. The authors also acknowledge the assistance of Dr Louise Owen and Dr C. Omar Kamlin at the Royal Women’s Hospital for advice on study implementation in the delivery room.

