## Supplementary material for "Unravelling the complexities of the first breaths of life": Online Supplementary Material

***Online Supplementary Materials***

1. Detailed Methods
2. Supplementary Results
3. Description of Supplementary Videos
4. Reference List for Online Supplementary Materials

**1. DETAILED METHODS**

***Study Population***

Recruitment was conducted in the operating theatres of The Royal Women’s Hospital, Melbourne, Australia (a tertiary obstetric hospital) between February to May 2017. The study was approved by the hospital’s Ethics in Human Research Committee, and registered with the Australian New Zealand Clinical Trials Registry (ACTRN12618000128291).

Infants were eligible for enrolment if they were delivered by elective caesarean section via spinal anaesthesia for non-fetal reasons at ≥36^+0^ weeks post menstrual age and there was a low likelihood of neonatal intervention in the delivery room. Potentially eligible infants were identified from elective surgical lists and written prospective consent was obtained from parents before delivery. Timing of cord clamping was at the discretion of the clinical team. Infants were not included if 1) the mother received any analgesia, sedation or anaesthesia that could suppress the infants respiratory behaviour at birth, 2) the placement of an electrical impedance tomography (EIT) belt in the standard position would interfere with clinical care,(E1-3) 3) the fetus was known to have a congenital condition or disease that may alter interpretability of the EIT data or 4) the clinical team requested the infant not be studied. If an enrolled infant received resuscitative interventions in the delivery room, data were recorded but excluded from analysis.

***Measurements***

Heart rate and peripheral oxygen saturation (SpO_2_) were displayed and measured with a Massimo Radical 7 pulse oximeter and newborn SET sensor (Massimo Corporation, Irvine CA). Regional lung volume changes were imaged with the Pioneer EIT system using 33 cm NeoSensor Belt and custom made neonatal connector system (Sentec AG, Landquart, Switzerland), containing a positional gyroscope placed in a supine position on an existing shelve of the neonatal resuscitaire.(E1-5) The Pioneer system is an open-source EIT platform designed to be modified for bespoke research purposes. We have previously used the system in the Neonatal Intensive Care Unit (E1, 3) and to image the respiratory transition in preterm lambs.(E4-11) The NeoSensor Belt consisted of a striped electrically conductive (Ag) non-adherent textile cover enclosing a 3D space fabric wrapped around a flexible printed circuit board containing 32 electrodes to allow unobstructed chest wall movement.(E2) Electrical conductance with the skin is achieved with warmed ultrasound gel applied to the belt before placement. EIT images were measured at 48 frames per second using a current injection amplitude of 3 mA_rms_ at a frequency of 200 kHz. Audio and video were recorded at 30 frames per second with a high-definition webcam (Logitech, Lausanne, Switzerland).

***Delivery room protocol***

Prior to birth the measurement equipment was placed on a neonatal resuscitaire and the study protocol reaffirmed with the clinical team caring for the infant. To ensure that the exact time of birth was digitally recorded in all systems, EIT and video/audio imaging recording commenced with the resuscitation clock. As the infant was being placed supine on the resuscitaire, the NeoSensor Belt was secured around the chest at nipple level, and below the armpit, using a Velcro tab, as shown in the **Supplementary Video 1**. We have used this system and method of attachment in preterm lambs, and refined the process in a pilot study (n=3 infants).(E4, 5, 8, 12) The pulse oximetry sensor was applied to the right hand. Apart from the application of the EIT belt and pulse oximetry sensor there was no interference with routine clinical care, and the infants were managed in a supine position in accordance with local guidelines. Data were recorded only during care on the resuscitaire. Measuring equipment was removed when the clinical staffed deemed the infant ready to be given to her/his parents, or transferred to another setting for ongoing clinical care.

The following time points were documented: the time the umbilical cord was cut, the infant was placed on the resuscitatire, the first breath, the EIT belt was applied (and duration of application), first EIT signal was acquired, pulse oximetry was applied and first oximetry signal acquired.

EIT remains a research tool in neonatal medicine (E13), and has never previously been used in the delivery room. Introduction of any new research technology into a clinical environment must only be done so without impacting clinical care or the patient wellbeing and experience. To allow staff to become familiar with the technology, and ensure that we did not impose of the unique mother-baby experience in the first few minutes after birth, we intentionally elected to limit the study population to infants receiving a period of separation from their mother as part of standard care (resuscitaire).

**Data acquisition and analysis**

Video, audio and pulse oximetry data were continuously recorded and analysed in LabChart (AD Instruments, Sydney, Australia). SpO_2_ and heart rate data was manually reviewed for loss of signal or movement artefact, and the mean and range of included data determined for each minute after birth. The following clinical data were collected: gestational age, birth weight, sex, Apgar score, need for stimulation and/or resuscitation after birth, reason for caesarean section and type of maternal anaesthesia. EIT data was recorded in real-time into the same computer (to ensure matched time-stamp signals) using a custom-built infant imaging package (Sentec AG) built using the EIDORS platform.(E14) This software provides a simple graphic interface to indicate electrode contact quality, real-time waveform of whole lung impedance change and a functional EIT image.

**Supplementary Figure 1** summarises the data analysis workflow. EIT data were only analysed if matched video and audio signals were present. Included time-course EIT images were reconstructed using the vendor-provided human model atlas with thoracic shape and lung and heart regions defined from a collection of computerised tomography images in the GREIT algorithm to minimise shape deformation.(E13, 15) Non-lung regions were excluded during post-processing and the pixel EIT signals filtered to the respiratory domain (IbeX software package, Sentec AG) as described previously.(E1, 3, 5, 11) Within the entire lung region (‘global’ signal), each potential tidal volume (V_T_) change due to breathing was identified. Analysis of the EIT change associated with a breath was only performed if there was 1) video confirmation of a breath (defined as chest wall or abdominal movement); and 2) there was no movement interference on the video (for example due to drying or wrapping), even if a breath was visualised. The audio signal was then reviewed for all included artefact-free and video-confirmed breaths and each classified by the presence of a cry, grunt or no breathing noise (tidal breath). If audio classification was not possible the breath was excluded.

For included breaths, the pre and post minima of the global impedance change was defined as the relative FRC before and after the breath. Inspiration time and subsequent relative V_T_ were defined as the time and the signal difference from the pre-inflation minima to the maxima, and expiration time and expiratory V_T_ change from the maxima to next minima. The pre and post-breath FRC, breath duration, inspiratory time (Ti), expiratory time (Te), time constant of the respiratory cycle (τ) and relative peak inspiratory (PIF) and expiratory flow (PEF) were calculated for the global signal and right, left, ventral and dorsal lung regions. The shape of the time-volume impedance change for each breath was visually reviewed by an investigator (DGT) blinded to the breath type and time from birth, and coded as consistent or not with volume change during tidal breathing (triangular shape with linear volume change during inspiration and expiration), to determine if breathing patterns during the respiratory transition differed from those seen in the already aerated lung.

To define the spatiotemporal distribution, and homogeneity, of V_T_ within the chest the centre of ventilation along the ventrodorsal (CoV_VD_) and right-left (CoV_RL_) planes were calculated and expressed as a percentage (0% representing all V_T_ in the most ventral or right and 100% in the most dorsal or left lung regions).(E13, 16) Due to the uneven contribution of each lung to the total lung volume, homogeneous ventilation occurs at CoV_VD_ 55% and CoV_RL_ 46%. The percentage of the global V_T_ signal was also calculated for the most dependent, central and non-dependent thirds of the right and left lung, and the percentage and location of lung regions without any apparent V_T_ signal determined.

**Sample size and statistical analysis**

In previous prospective observational studies of respiratory parameters after birth approximately 40% of studied infants needed to be excluded for technical reasons.(E17) It was also estimated that 5% of infants would require resuscitation in the delivery room. A convenience sample of thirty infants was required to provide data for breath-by-breath classification and analysis of 15-20 infants. A median of 62 inflations/infant over 90s were recorded in a study of facemask flow, volume and pressure characteristics (without video/audio) in a study of 13 term infants immediately after birth (the previous largest study of breath-by-breath characteristics at birth).(E17) As our study included stricter analysis criteria we anticipated 20-30 breaths/minute/infant would be included for breath-by-breath classification and analysis (100-150 breaths per 5-6 minute period per infant).

Data were analysed for each minute after birth (up to a maximum of 6 minutes). All data were analysed for outliers using a 1% threshold (ROUT). Continuous data were tested for normality and analysed with a mixed-effects linear regression model, with robust standard error and cluster analysis to adjust for multiple breaths from each infant if appropriate. Due to the variable end point, breaths during the fifth and sixth minute (241-360s) of the study were pooled for statistical analysis. Statistical analysis was performed using Prism (v8.2.1, GraphPad Software, San Diego, CA) or Stata (v16.0, StataCorp, College Station, TX), and a p value <0.05 considered statistically significant.

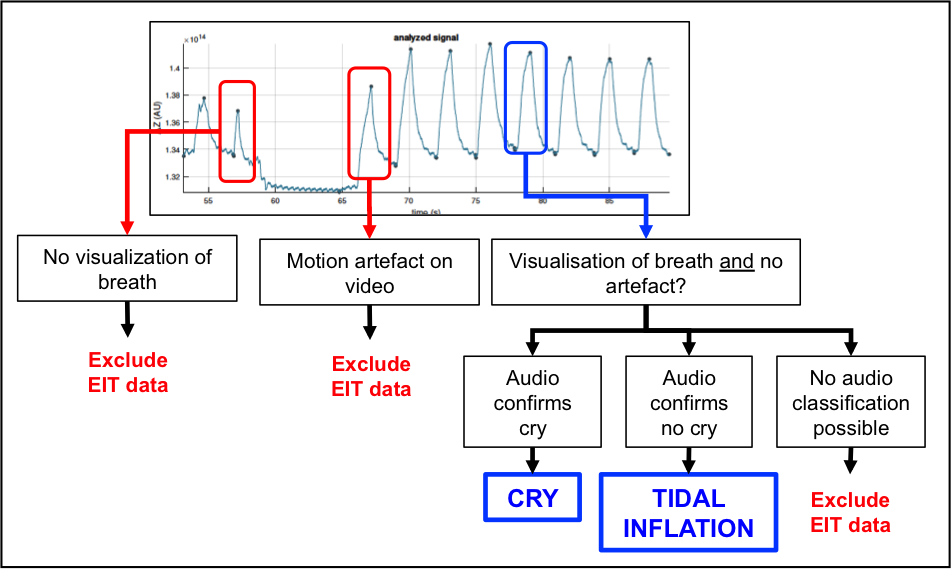

**Supplementary Figure 1.** **Data analysis workflow.** Simultaneous video and audio recordings were made with EIT imaging once the infant was placed on the resuscitaire as demonstrated in **Supplementary Videos 1 and 2**. Time-course EIT data was only included for breath-by-breath analysis if matched signals for EIT, video and audio were obtained. Volume changes in the global time-course EIT signal (IbeX Software package, Sentec AG, Landquart, Switzerland) were identified (red and blue boxes). Any EIT volume change was only classified as a breath if there was visual confirmation of a breath on video (chest wall and/or abdominal movement) and no movement artefact (for example handling). Finally, each video confirmed breath on EIT was then classified as a cry, grunt or tidal breath based on the noise, or lack thereof, on audio signal. If no audio classification was possible (for example too much background noise) the breath was excluded from analysis.

**2. RESULTS**

Presented in order mentioned in main manuscript results section.

**Online Supplemental Material Table 1. Infant Characteristics (n=17)**

| **GA (weeks)** | **BW (kg)** | | **Gender (F:M)** | **DCC** | **Cried at birth** | **Stimulation at birth** | **Apgar Score** | |
| --- | --- | --- | --- | --- | --- | --- | --- | --- |
|  |  |  |  |  |  |  | **1 min** | **5 min** |
| 39 (36, 39) | 3.26 (2.77, 3.86) | | 7:10 | 2 (11)%* | 15 (88%) | 8 (47%) | 9 (7,9) | 9 (9,9) |
| **Anaesthesia** | | **Reason for caesarean section** | | | | | | |
| 16 spinal; 1 general anaesthesia | | 16 previous caesarean section (3 gestational diabetes, 1 maternal muscular dystrophy, 1 breech), 1 suspected macrosomia, 1 previous stillbirth | | | | | | |

*Abbreviations:* GA; Gestational age at birth (completed weeks), BW; birth weight, DCC; Delayed Cord Clamping.

*Umbilical cord was cut at 30 and 40 s. All data median (range) unless stated.

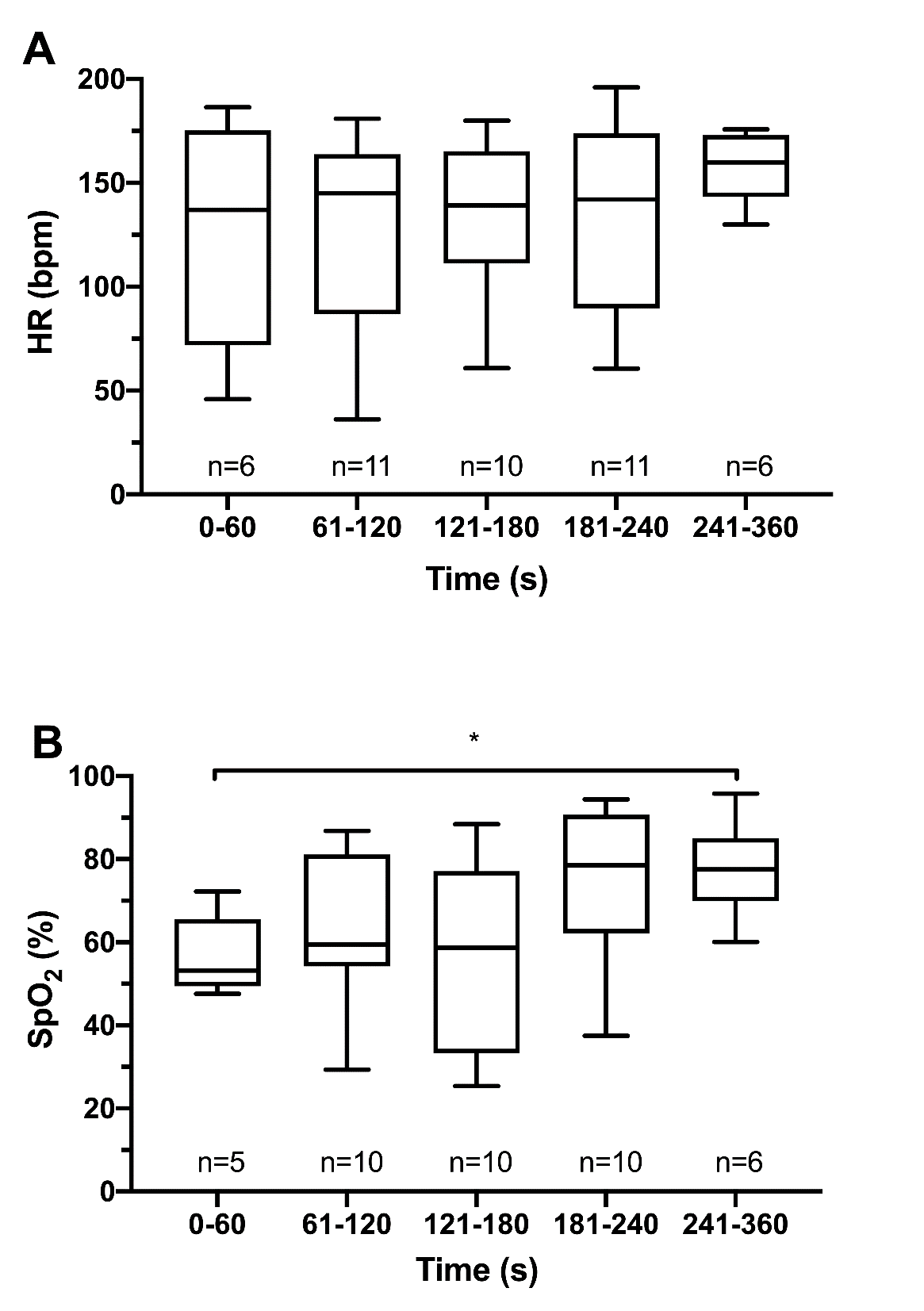

**Supplementary Figure 2.** Heart rate (HR; **A**) and peripheral saturation of oxygen (SpO_2_; **B**) for each minute from birth until six minutes after birth. Boxes median and 10-90^th^ percentile and error bars range. Numbers below each box represent the number of infants with complete SpO_2_ or heart rate data for that minute. *p=0.029 (mixed effects model).

**Regional breathing patterns**

***Inspiratory Time***

Ti behaved similarly in the ventral (p=0.25, mixed-effects model) and dorsal (p=0.53) lung during crying (Supplementary Figure 3). During tidal breaths the Ti was variable in the ventral lung over time (p=0.003), but not the dorsal lung (p=0.56). In the ventral lung Ti during tidal breaths was a mean (95% CI) 140 (55, 245) ms longer after 240s compared to 121-240s. There was no difference in the ventrodorsal behaviour of Ti at each time epoch for either crying or tidal breaths. After 120s, Ti was shorter during crying in the ventral and dorsal lungs, with the greatest difference being 138 (38, 239) ms and 70 (19, 121) ms respectively after 240s.

Both the right and left lung Ti was the same over time during crying and tidal breaths. Ti was shorter during crying compared to tidal breaths after 121s in both the right and left lung; mean (95% CI) difference for the right lung at -112 (14, 209) ms (121-180s) and -125 (61, 189) ms (after 180s), with similar differences in the left lung.

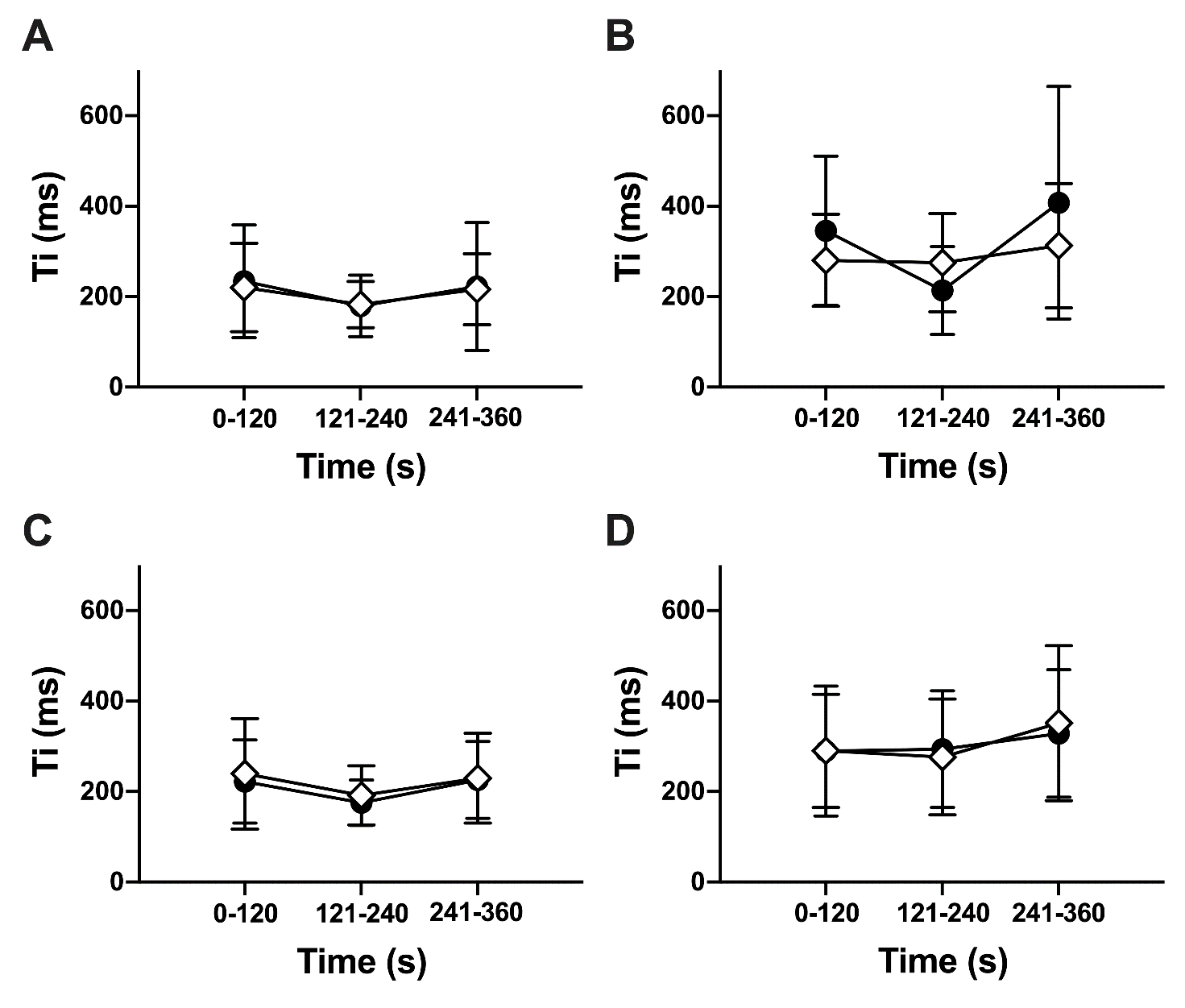

**Supplementary Figure 3.** Spatiotemporal behaviour of Ti during crying (**A** and **C**) and tidal (**B** and **D**) breaths along the ventrodorsal (**A** and **B**) and right-left (**C** and **D**) lung planes. Black circles represent right or ventral lung respectively, and open diamonds left or dorsal lung. All data mean±SD. *p<0.05, **p<0.01, ***p<0.0001 between lung regions at time point (mixed-effects model).

***Expiratory Time***

Te increased with time in the dorsal lung (p=0.007; mixed-effects model) during crying, but did not change in the ventral lung (Supplementary Figure 4). After 180s Te was a mean (95% CI) 234 (102, 367) ms longer in the dorsal lung. Te did not change in the ventral (p=0.73) or dorsal (p=0.49) lung over time, or between regions. The type of breath did not influence Te at any time epoch within the ventral and dorsal lung.

During crying Te did not change with time in the right (p=0.19; mixed-effects model) and left (p=0.35) lungs during cries, and both the right and left lung Te were similar. Te was unchanged with time in the right lung during tidal breaths (p=0.12), but decreased in the left lung, being a mean (95% CI) 179 (6, 353) ms shorter after 240s compared to the first two minutes. Tidal breath Te was not different between the right and left lung at each time epoch.

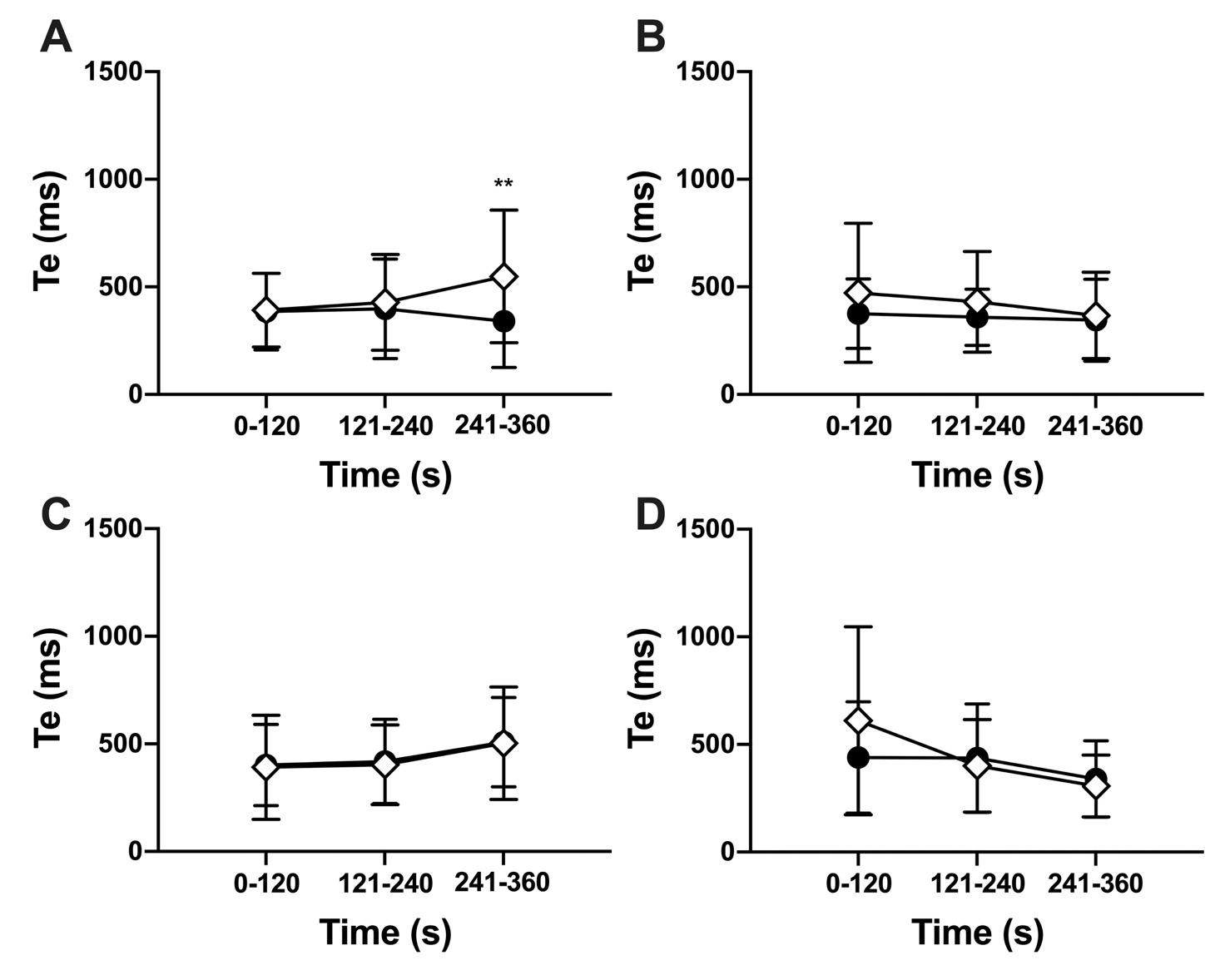

**Supplementary Figure 4.** Spatiotemporal behaviour of Te during crying (**A** and **C**) and tidal (**B** and **D**) breaths along the ventrodorsal (**A** and **B**) and right-left (**C** and **D**) lung planes. Symbols as per OSM Figure 3. All data mean±SD.

***Respiratory system time constant***

The respiratory time constant was a mean (95% CI) 111 (3, 192) ms longer after 240s compared to 121-240s in the ventral lung during crying (mixed-effects model; Supplementary Figure 5). There was no difference in τ with time in the dorsal lung (p=0.94) during crying, and both the ventral (p=0.75) and dorsal (p=0.71) lungs during tidal breaths. Cry and tidal breath τ were the same within the ventral (p=0.60) and dorsal (p=0.77) lung.

The right and left lung τ was similar, and did not change with time during both cries (p=0.69 and p=0.50 respectively; mixed-effects model) and tidal breath (p=0.20 and p=0.41). There was no difference in τ in the right (p=0.26) and left (p=0.37) lung between cries and tidal breaths. During crying the ventral lung PIF was greater at all time epochs than in the dorsal lung (p<0.0001), with the greatest difference of 0.95 (0.81, 1.10) AU/s occurring between 121-240s.

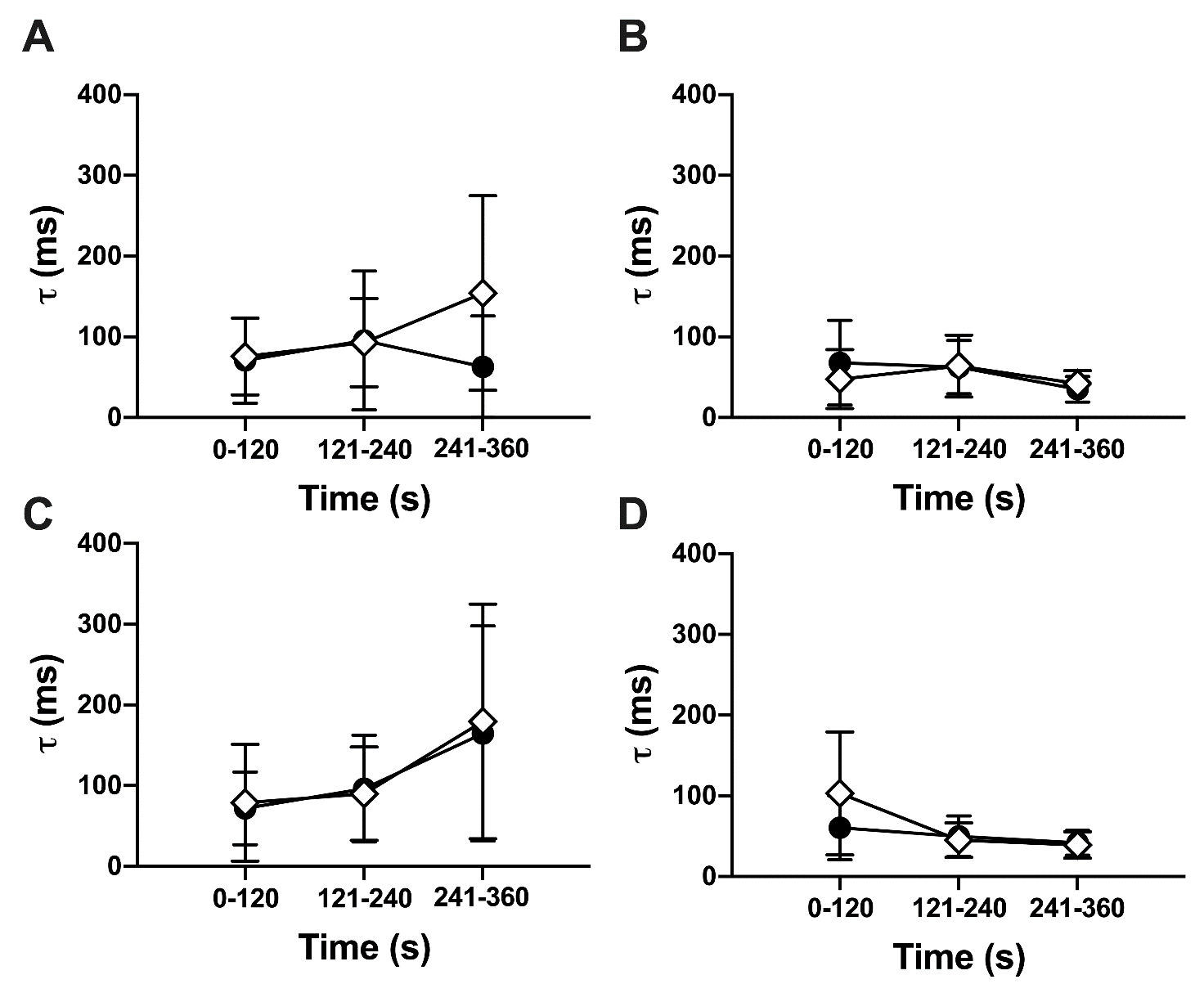

**Supplementary Figure 5.** Spatiotemporal behaviour of τ during crying (**A** and **C**) and tidal (**B** and **D**) breaths along the ventrodorsal (**A** and **B**) and right-left (**C** and **D**) lung planes. Symbols as per OSM Figure 3. All data mean±SD.

***Peak Inspiratory Flow***

PIF did not change over time in the ventral and dorsal lung during crying (Supplementary Figure 6), but was always faster in the dorsal lung at all time epochs (all p<0.0001). During tidal breaths the ventral lung PIF between 121-240s was a mean (95% CI) 0.97 (0.61, 1.33) AU/s and 1.13 (0.90, 1.35) AU/s faster than during the first two minutes and after 240s respectively (mixed-effects model). The dorsal lung PIF did not change over time during tidal breaths. During tidal breaths the ventral and dorsal lung PIF only differed after 240s; mean (95% CI) difference 0.34 (0.28, 0.39) AU/s. Within both the ventral and dorsal lung the PIF was significantly faster during crying than tidal breaths at each time epoch except ventral lung at 121-240s (both p<0.0001).

PIF varied with time in the right and left lung during crying, being a mean (95% CI) 0.76 (0.42, 1.10) AU/s greater between 121-240s and after 240s in the right lung, and 0.49 (0.29, 0.69) AU/s between 121-240s and 0-120s in the left lung (mixed-effects model). PIF decreased with time in the right lung (p<0.0001) during tidal breaths, and was 0.52 (0.35, 0.70) AU/s slower after 240s compared to 121-240s in the left lung. During crying PIF was significantly greater at all time epochs in the right lung compared to the left lung (p<0.0001), with the greatest difference of 1.67 (1.38, 1.96) AU/s occurring between 0-120s. PIF was only differed within the right and left lung between 0-120s during tidal breaths; 1.06 (0.51, 1.61) AU/s. Within the right and left lung, the PIF was greater at all time epochs during crying compared to tidal breaths (both p<0.0001), with the differences being greater in the right lung.

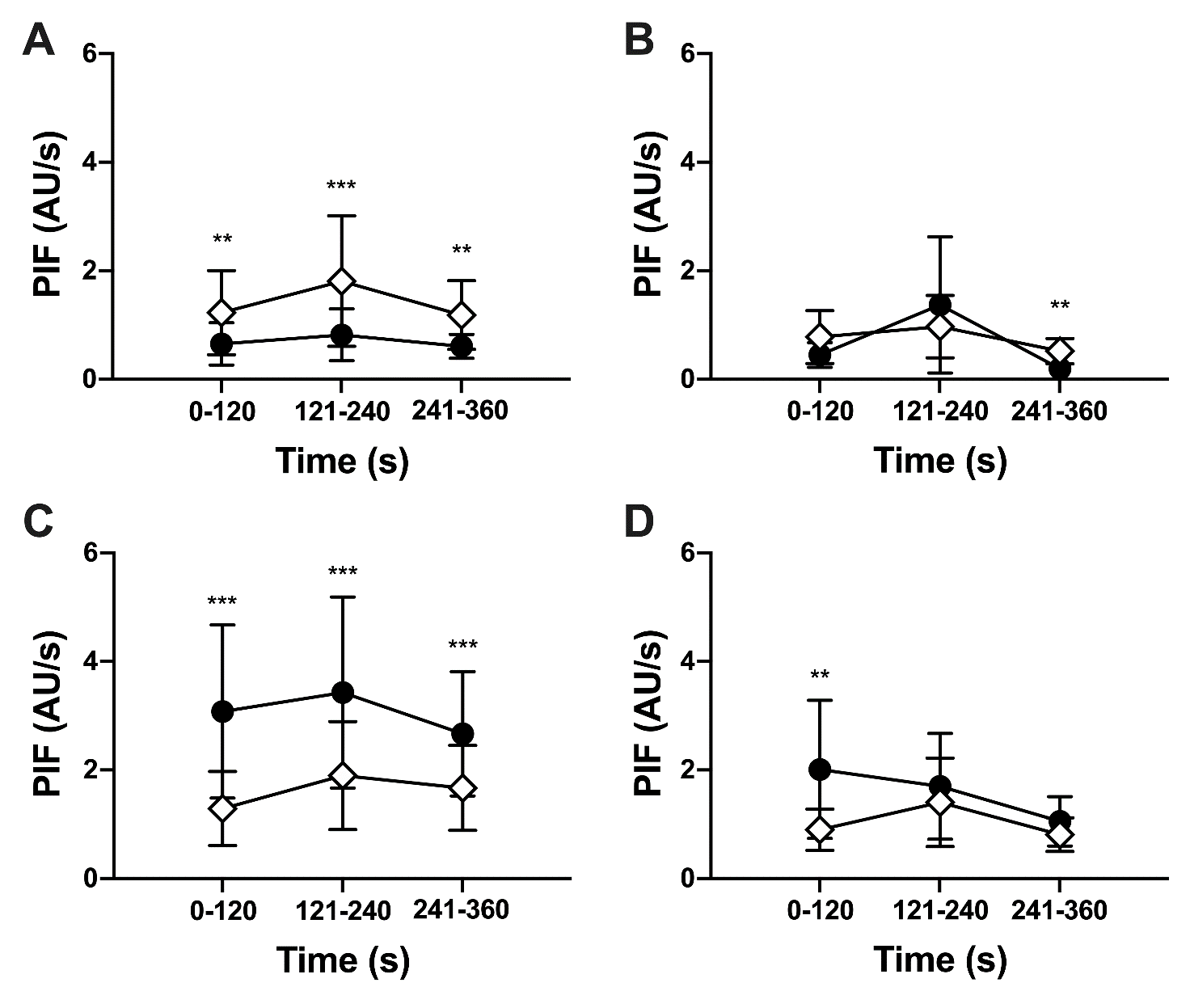

**Supplementary Figure 6.** Spatiotemporal behaviour of PIF during crying (**A** and **C**) and tidal (**B** and **D**) breaths along the ventrodorsal (**A** and **B**) and right-left (**C** and **D**) lung planes. Symbols as per OSM Figure 3. All data mean±SD.

Within the ventral lung crying and tidal breaths differed by a mean (95% CI) 0.58 (0.34, 0.82) AU/s, -0.40 (-0.63, -0.17) AU/s and 0.32 (0.27, 0.36) AU/s at 0-120s, 121-240s and after 241s respectively (mixed-effects model). Within the dorsal lung PIF was 0.57 (0.30, 0.85) AU/s, 0.79 (0.53, 1.06) AU/s and 0.55 (0.42, 0.68) AU/s greater during crying than tidal breaths at 0-120s, 121-240s and after 241s respectively. During tidal breath PIF was a 0.82 (0.55, 1.08) AU/s greater between 0-120s compared to after 240s in the right lung. Within the right lung crying resulted in a 1.07 (0.62, 1.52) AU/s, 1.48 (0.98, 1.97) AU/s and 1.46 (1.23, 1.72) AU/s greater PIF than tidal breaths at 0-120s, 121-240s and after 240s. PIF was also greater in the left lung during crying; 0.46 (0.14, 0.77) AU/s, 0.50 (0.23, 0.77) AU/s and 0.83 (0.68, 0.98) AU/s.

***Peak Expiratory Flow***

PEF did not change with time in the ventral lung (p=0.12; mixed-effects model) but increased in the dorsal lung (p=0.0008) with crying (Supplementary Figure 7). PEF was similar over time in the ventral and dorsal lung. During crying PEF was always greater in the ventral lung, with a mean (95% CI) difference of 0.21 (0.12, 0.30) AU/s, 0.49 (0.42, 0.55) AU/s and 0.42 (0.32, 0.52) AU/s in each time epoch. During tidal breaths the ventral and dorsal lung PEF was only different after 240s; 0.32 (0.25, 0.38). Within the ventral lung crying resulted in greater PEF during the first two minutes and after 240s; 0.23 (0.09, 0.36) AU/s and 0.20 (0.16, 0.25) AU/s. PEF was greater during crying within the dorsal lung at all time points, increasing from a 0.27 (0.12, 0.42) AU/s difference in the first two minutes to 0.31 (0.22, 0.39) AU/s after 240s.

PEF was greater in the right lung for both crying and tidal breaths (p<0.0001; mixed-effects model). During crying the right lung PEF did not change with time (p=0.06) whilst PEF increased with time in the left lung (p<0.0001). The right lung PEF decreased with time during tidal breaths (p=0.039), and left lung increased (p<0.0001). In the first two minutes PEF was a mean (95% CI) -0.78 (-1.00, -0.56) AU/s greater in the right than left lung for cries, and -0.71 (-1.04, -0.37) AU/s for tidal breaths. After 240s these differences were -0.46 (-0.61, -0.31) AU/s and -0.27 (-0.41, -0.13) AU/s respectively. Within the right lung PEF was greater at all time points during cries compared to tidal breaths (p<0.0001). PEF was only different between cries and tidal breaths after 240s in the left lung; -0.40 (-0.53, -0.27) AU/s.

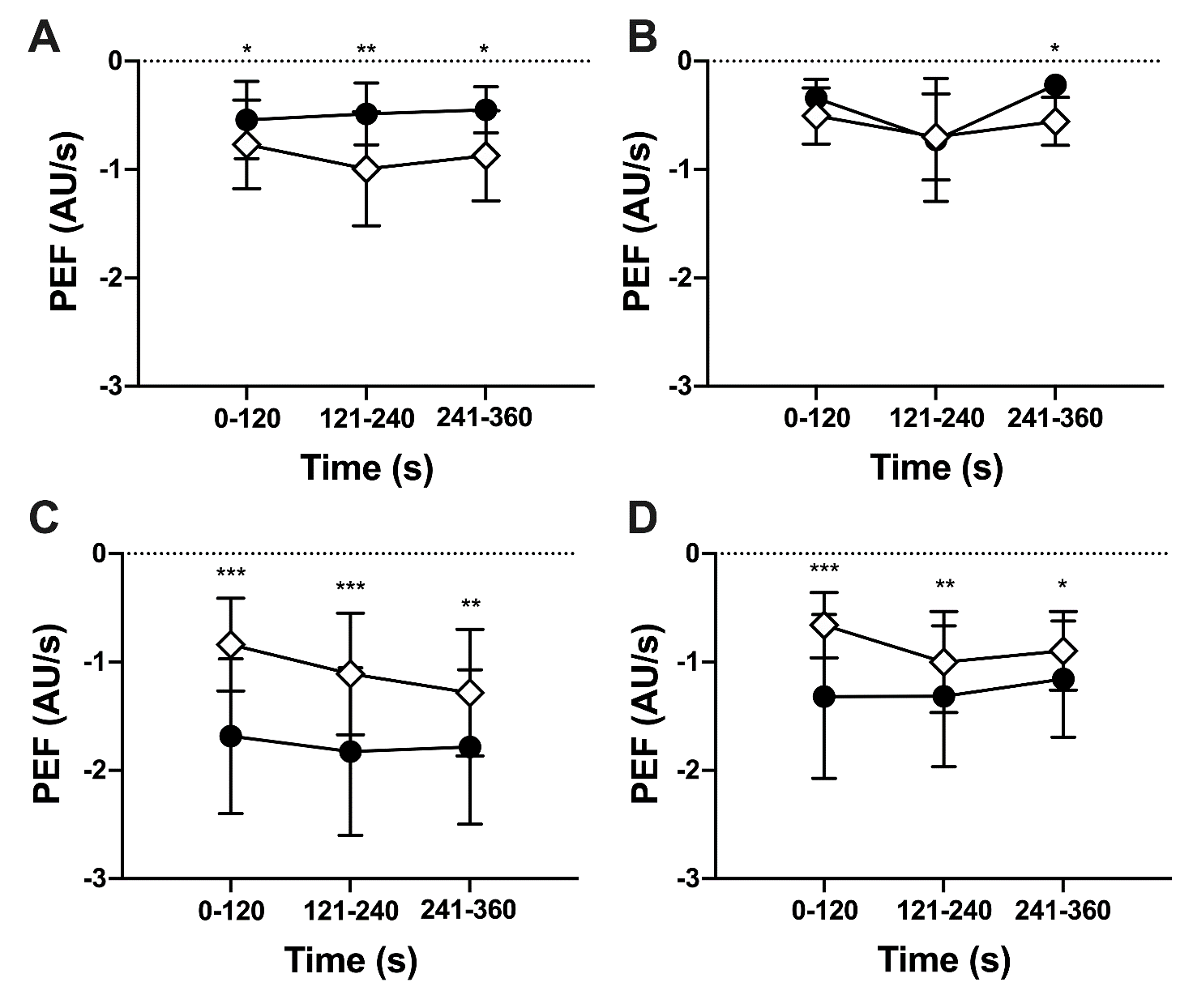

**Supplementary Figure 7.** Spatiotemporal behaviour of PEF during crying (**A** and **C**) and tidal (**B** and **D**) breaths along the ventrodorsal (**A** and **B**) and right-left (**C** and **D**) lung planes. Symbols as per OSM Figure 3. All data mean±SD.

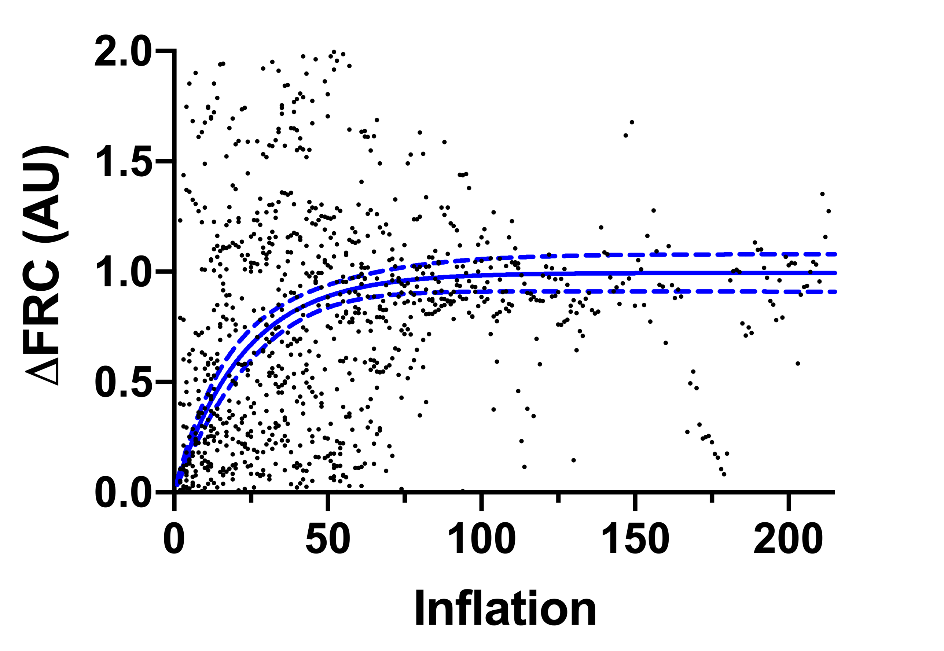

**Supplementary Figure 8.** Change in absolute EIT values (AU; arbitrary units) from first measured breath (functional residual capacity; FRC) for all breaths. Blue line represents line of best fit (dashed lines 96% CI) using a one-phase exponential association; y=y_plateau_.[1-*e*^x.τ-1^]; plateau (95% CI) 0.99 (0.91, 1.09) AU, τ 22.3 (16.6, 29.4) breaths (R^2^ 0.08, RSME 0.83, replicates test discrepancy (F) 0.20 [p>0.99]).

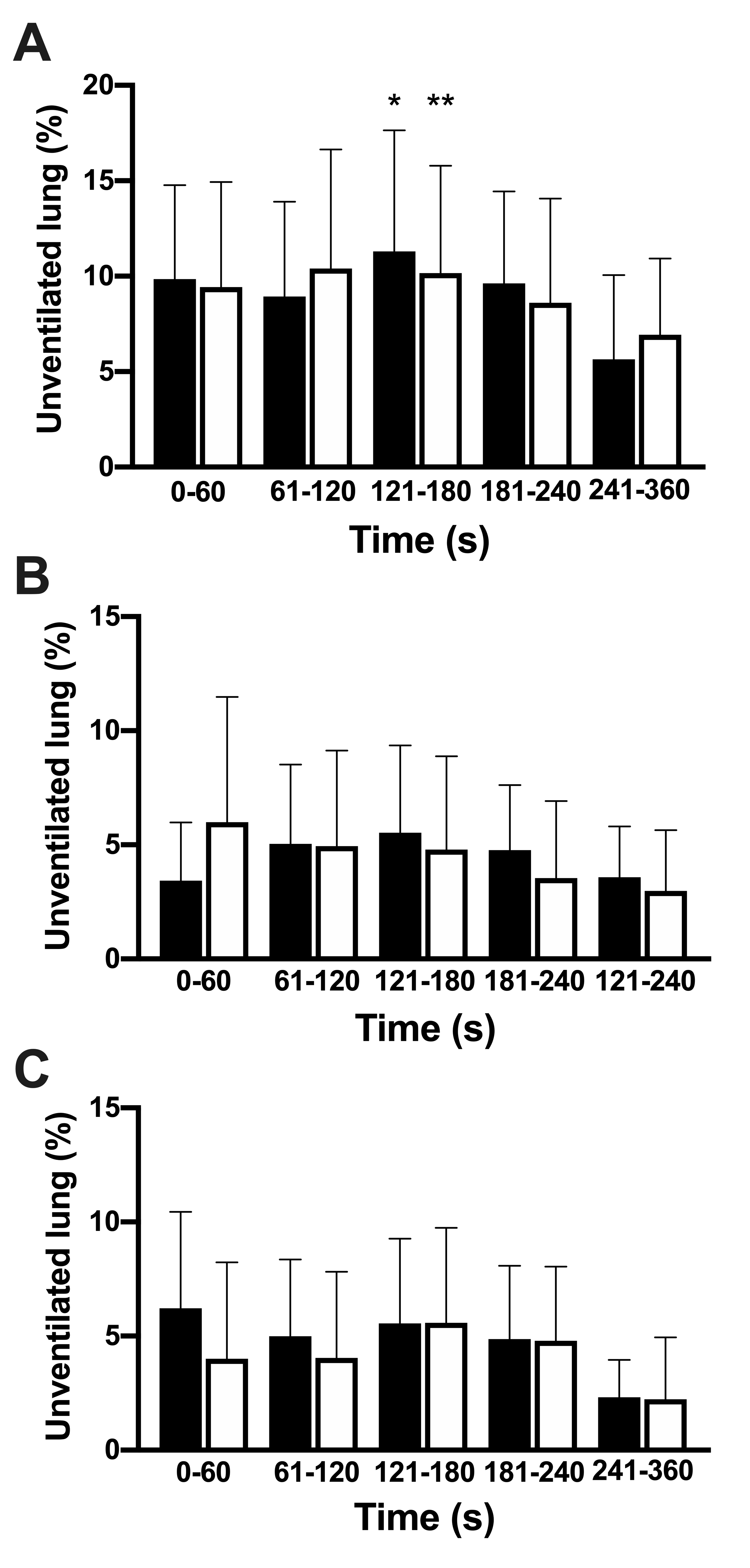

**Supplementary Figure 9. A.** Percentage of lung regions without any apparent ventilation for crying (black) and tidal (white) breaths by minute from birth. Only tidal breaths had a significant change in unventilated lung over time; p=0.003 (tidal) versus p=0.11 (cry); mixed-effects model. 121-180s vs 241-360s: * p=0.027; mean (95% CI) 3.3 (0.4, 6.1)%, **p<0.01; 4.3 (1.2, 7.5)%. Percentage of gravity non-dependent (black bars) and dependent (white bar) lung regions without any apparent ventilation for crying (**B**) and tidal (**C**) breaths by minute from birth. All data mean±SD.

**3. DESCRIPTION OF SUPPLEMENTARY VIDEOS**

**Supplementary Video 1.** Video footage and audio of the experimental methodology being applied to a representative infant during normal clinical care on the resuscitaire immediately after delivery. Video shows application of the EIT belt (chest) and pulse oximetry sensor (left hand). Video and audio delineate periods of crying and tidal breathing. The infants face and other identifiable features of participants in the video have been obscured. Written parental permission was obtained for inclusion of video footage and audio in this publication. Video recording was made at the Royal Women’s Hospital (Melbourne, Victoria) on the 19^th^ of May 2017. Video duration 05:07 min:sec.

**Supplementary Video 2.** Video abstract of the key findings, and differences, between crying and tidal breathing at birth and our rationale explaining the importance of these events in the respiratory transition to air-breathing at birth. Video duration 06:15 min:sec.

E4. Tingay DG, Togo A, Pereira-Fantini PM, Miedema M, McCall KE, Perkins EJ, Thomson J, Dowse G, Dellaca RL, Davis PG, Dargaville PA. Aeration strategy at birth influences the physiological response to surfactant in preterm lambs. Arch Dis Child Fetal Neonatal Ed 2019.

E5. Tingay DG, Pereira-Fantini PM, Oakley R, McCall KE, Perkins EJ, Miedema M, Sourial M, Thomson J, Waldmann A, Dellaca RL, Davis PG, Dargaville PA. Gradual Aeration at Birth is More Lung Protective than a Sustained Inflation in Preterm Lambs. Am J Respir Crit Care Med 2019; 200: 609-616.

E6. Tingay DG, Rajapaksa A, Zannin E, Pereira-Fantini PM, Dellaca RL, Perkins EJ, Zonneveld CE, Adler A, Black D, Frerichs I, Lavizzari A, Sourial M, Grychtol B, Mosca F, Davis PG. Effectiveness of individualized lung recruitment strategies at birth: an experimental study in preterm lambs. Am J Physiol Lung Cell Mol Physiol 2017; 312: L32-L41.

E7. Milesi I, Tingay DG, Lavizzari A, Bianco F, Zannin E, Tagliabue P, Mosca F, Ventura ML, Rajapaksa A, Perkins EJ, Black D, Di Castri M, Sourial MD, Pohlmann G, Dellaca RL. Supraglottic Atomization of Surfactant in Spontaneously Breathing Lambs Receiving Continuous Positive Airway Pressure. Pediatr Crit Care Med 2017.

E8. McCall KE, Waldmann AD, Pereira-Fantini P, Oakley R, Miedema M, Perkins EJ, Davis PG, Dargaville PA, Bohm SH, Dellaca R, Sourial M, Zannin E, Rajapaksa AE, Tan A, Adler A, Frerichs I, Tingay DG. Time to lung aeration during a sustained inflation at birth is influenced by gestation in lambs. Pediatr Res 2017; 82: 712-720.

E9. Tingay DG, Rajapaksa A, Zonneveld CE, Black D, Perkins EJ, Adler A, Grychtol B, Lavizzari A, Frerichs I, Zahra VA, Davis PG. Spatiotemporal Aeration and Lung Injury Patterns Are Influenced by the First Inflation Strategy at Birth. Am J Respir Cell Mol Biol 2016; 54: 263-272.

E10. Tingay DG, Lavizzari A, Zonneveld CE, Rajapaksa A, Zannin E, Perkins E, Black D, Sourial M, Dellaca RL, Mosca F, Adler A, Grychtol B, Frerichs I, Davis PG. An individualized approach to sustained inflation duration at birth improves outcomes in newborn preterm lambs. Am J Physiol Lung Cell Mol Physiol 2015; 309: L1138-1149.

E11. Tingay DG, Bhatia R, Schmolzer GM, Wallace MJ, Zahra VA, Davis PG. Effect of sustained inflation vs. stepwise PEEP strategy at birth on gas exchange and lung mechanics in preterm lambs. Pediatr Res 2014; 75: 288-294.

E12. Miedema M, McCall KE, Perkins EJ, Oakley RB, Pereira-Fantini PM, Rajapaksa AE, Waldmann AD, Tingay DG, van Kaam AH. Lung Recruitment Strategies During High Frequency Oscillatory Ventilation in Preterm Lambs. Front Pediatr 2018; 6: 436.

E13. Frerichs I, Amato MB, van Kaam AH, Tingay DG, Zhao Z, Grychtol B, Bodenstein M, Gagnon H, Bohm SH, Teschner E, Stenqvist O, Mauri T, Torsani V, Camporota L, Schibler A, Wolf GK, Gommers D, Leonhardt S, Adler A. Chest electrical impedance tomography examination, data analysis, terminology, clinical use and recommendations: consensus statement of the TRanslational EIT developmeNt stuDy group. Thorax 2017; 72: 83-93.

E14. Adler A, Lionheart WR. Uses and abuses of EIDORS: an extensible software base for EIT. Physiol Meas 2006; 27: S25-42.

E15. Adler A, Arnold JH, Bayford R, Borsic A, Brown B, Dixon P, Faes TJ, Frerichs I, Gagnon H, Garber Y, Grychtol B, Hahn G, Lionheart WR, Malik A, Patterson RP, Stocks J, Tizzard A, Weiler N, Wolf GK. GREIT: a unified approach to 2D linear EIT reconstruction of lung images. Physiol Meas 2009; 30: S35-55.

E16. Frerichs I, Becher T. Chest electrical impedance tomography measures in neonatology and paediatrics-a survey on clinical usefulness. Physiol Meas 2019; 40: 054001.

E17. te Pas AB, Wong C, Kamlin CO, Dawson JA, Morley CJ, Davis PG. Breathing patterns in preterm and term infants immediately after birth. Pediatr Res 2009; 65: 352-356.
